# Tracking Student Wellbeing Throughout the COVID-19 Pandemic

**DOI:** 10.1101/2025.08.29.25334729

**Authors:** Donald J. Noble, Charles L. Raison

**Affiliations:** The Center for the Study of Human Health, Emory University, Atlanta, GA, USA

**Keywords:** COVID-19 pandemic, mental wellbeing, observational study, social isolation, undergraduate health

## Abstract

**Background:** The COVID-19 pandemic served as a global, uncontrolled social isolation experiment, with especially pernicious effects on mental health in young adults. We sought to understand how the COVID-19 pandemic and associated social restrictions impacted the wellbeing of university undergraduate students.

**Methods:** 277 total U.S. undergraduate students enrolled in a course on mental wellbeing and resilience that was offered once each year from 2020 to 2024. Students anonymously completed surveys of anxiety, depression, and wellbeing on a weekly basis. These behavioral data were aggregated and investigated for associations with local COVID-19 case levels and a university social gathering meter.

**Results:** Average student wellbeing declined a few months into the COVID-19 pandemic in Fall 2020, remaining low in Fall 2021 and Spring 2022, with 63.7% of students at-risk for poor wellbeing based on standard cut-offs over these three semesters. Depression and anxiety peaked during Fall 2021 with 39.0% and 34.5% of students at-risk for anxiety and depressive disorders, respectively. Mental health gradually improved following the return to in-person learning in mid-Spring 2022. Over all five semesters, survey questions reflecting anhedonia strongly associated with social gathering restrictions whereas questions assessing acute anxiety most strongly associated with local COVID-19 case levels.

**Conclusions:** Our findings highlight the negative impact of the COVID-19 pandemic on university student wellbeing and suggest partially separable influences of COVID-19 infection prevalence and social isolation on levels of student anxiety and anhedonia. More research is needed to minimize the mental costs of future pandemics in an increasingly interconnected world.

## 1 Introduction

The COVID-19 pandemic served as a global, uncontrolled social isolation experiment, with lasting consequences for physical and mental health. This was especially true in adolescents and young adults. A rapid systematic review published near the onset of the pandemic warned of damaging depressogenic and anxiogenic effects of social isolation and loneliness in children and adolescents, especially when prolonged (1), consistent with the effects of social isolation and loneliness on mental health being more debilitating in young vs. older adults (2).

Undergraduate students are a particularly vulnerable population, given that many of them have left home for the first time and are therefore particularly dependent on the types of close peer contact limited by social restriction measures during COVID-19. Negative impacts of the pandemic and associated social restrictions on student mental health have been observed globally, including in Italy (3), France (4), China (5), and Germany, where 72.2% of undergraduate respondents endorsed serious impairments in wellbeing (6). American students similarly reported harmful effects of decreased social interactions due to physical distancing during COVID-19 closures (7).

Social isolation alone likely does not account for the entire mental health burden of COVID-19. In the United States, by the third quarter of 2022, approximately two-thirds of the population had been infected with the virus (8). A large body of research demonstrates that the types of inflammatory processes engendered by the virus can cause depression (9). Completing a vicious cycle, inflammatory stimuli can also induce feelings of social isolation (10), and inflammation activates brain regions that mediate feelings of social disconnection (11). While social distancing reduced risk of death from COVID-19, at least some of this benefit was likely counterbalanced by the fact that mortality due to loneliness and social isolation is comparable to that from well-established risk factors like smoking and greater than risks due to obesity and hypertension (12). Given that young people were at reduced risk of COVID-19 mortality, the trade-offs between viral protection and the mental and physical health costs of isolation were likely especially unfavorable.

As part of a quality improvement project for our elective course titled “Mental Wellbeing and Resilience”, we obtained data on student wellbeing over three ‘COVID’ semesters (Fall [F] 2020-2021 and Spring [S] 2022) and two ‘recovery’ semesters (S23-24). Student mental health was measured using established surveys for anxiety (7-item Generalized Anxiety Disorder Scale [GAD-7]), depression (2-Item Patient Health Questionnaire [PHQ-2]), and general wellbeing (World Health Organization-Five Wellbeing Index [WHO5]). Fortuitously, data collection aligned with COVID-induced social isolation and recovery measures. Masking and social distancing requirements peaked in F20 and were gradually rescinded at the university level in F21, with a full return to largely mask-free in-person classes and social gatherings in late S22. Even as our course remained online all five semesters, the large majority of each student’s on-campus experience reflected this gradual lifting of pandemic restrictions. Here we present findings on the interrelationship between COVID-19 levels, social restriction measures, and student wellbeing.

## 2 Methods

### Study Overview

Data that comprise the current study were initially collected for course quality improvement purposes. Because of this, the study was not pre-registered. Recognizing upon completion of data collection that results were highly relevant to the impact of COVID-19 on student wellbeing, study authors petitioned the Emory University Institutional Review Board (IRB) and the study was reviewed and deemed exempt from IRB approval. The authors assert that all procedures contributing to this work comply with the ethical standards of the relevant national and institutional committees on human experimentation and with the Helsinki Declaration of 1975, as revised in 2013.

### Participants

277 Emory University undergraduate students who enrolled in the course titled “Mental Wellbeing & Resilience” over five semesters from 2020 to 2024 were included in this longitudinal observational study. The course was offered once per year, in the spring (2022-24) or fall (2020-21). The breakdown of different class years (Freshmen through Seniors) is reported for each semester in **Table S1** in the Supplementary Material. Students were notified in class by course instructors prior to the end of the enrollment period that their enrollment in the course constituted an agreement to anonymously fill out the weekly surveys. Links to university and external mental health resources and contact information were provided on Canvas and mentioned once per week in class. School credit (1 class participation point) was awarded for completion of each survey.

### Anxiety, Depression and Wellbeing Scales and Percent At-Risk

Students anonymously answered the clinically validated GAD-7, PHQ-2 and WHO5 each week throughout the 13-14 week course. Survey questions, answer options, and recommended cutoffs for further mental health screening are shown in **Table S2** in the Supplementary Material. Total possible scores on the GAD-7 ranged from 0 to 21 (higher = more anxious), on the PHQ-2 from 0 to 6 (higher = more depressed), and on the WHO5 from 0 to 25 (higher = greater wellbeing). Responses were submitted online through the Canvas Learning Management System, via Likert-type scales. Surveys were typically open for 5-7 days each week and closed on Wednesday night (Fall 2021, Spring 2022, Spring 2024), Thursday morning (Fall 2020), or Monday night (Spring 2023). While we did not directly screen students for DSM-5 depressive or anxiety disorders, each of the scales provides recommendations on the thresholds for further clinical screening. We calculated the metric *percent at-risk* as the percentage of weekly responses that reached these criteria for each survey.

### COVID-19 Cases and Social Gathering Meter

COVID-19 county level case numbers from DeKalb, Georgia were obtained from a publicly available New York Times /CDC database (https://github.com/nytimes/covid-19-data). COVID-19 case levels in university students, faculty, and staff were obtained from a publicly available dashboard with assistance from Emory’s UIT Data Solutions Service Center. All data were deidentified prior to study access and analysis. Weekly COVID-19 case numbers were calculated by summing total cases in the seven days preceding each survey deadline. For the social gathering meter, study investigators reviewed publicly available university announcements and assembled relevant guidelines into a color-coded timeline of key events reflecting tightened or loosened campus social gathering restrictions.

### Data Analysis

Each semester represented a unique cohort of individuals, whereas weekly scores within a semester generally included the same pool of students (with the exception of those who changed enrollment over the first few weeks of each semester or failed to submit complete surveys on a given week). Analyses were performed separately for each scale since they purport to measure different aspects of overall wellbeing and because they have different maximal scores. Nearly all study subjects provided scores at each week, leading to similar within-week variabilities, but the anonymous nature of the data precluded us from performing repeated-measures analysis on a within-subjects basis. Weekly survey scores (totals and percent at-risk) were organized by semester and averaged across weeks to derive overall mean and standard deviation values for each semester. Means were then compared using one-way ANOVA, treating semester as a fixed factor. In the case of significant results, we performed post-hoc testing to assess differences between semesters using Tukey’s test. To understand potential predictors of changes in wellbeing, correlation analyses with subsequent linear regression were performed for each scale with preselected variables considered to be the best representation of social isolation (university gathering meter levels) and local COVID-19 prevalence (COVID-19 case numbers in Dekalb County and university students, faculty, and staff). Correlations were performed using weekly data, rather than semesterly averages. Data are presented as mean ± SD unless otherwise indicated, with two-tailed tests and significance set at *P* ≤.05.

## 3 Results

### 3.1. Percent At-risk for Anxiety, Depression, and Wellbeing Scales Over Five Years

We sought to understand how weekly wellness levels assessed through the three surveys (GAD-7, PHQ-2, and WHO5) changed over five semesters (**Figure 1**). Week-by-week trajectories for survey means and percent at-risk are shown in **Figure 1A**. Mean scores are reported as Results S1 in the Supplementary Material.

**Figure 1.**
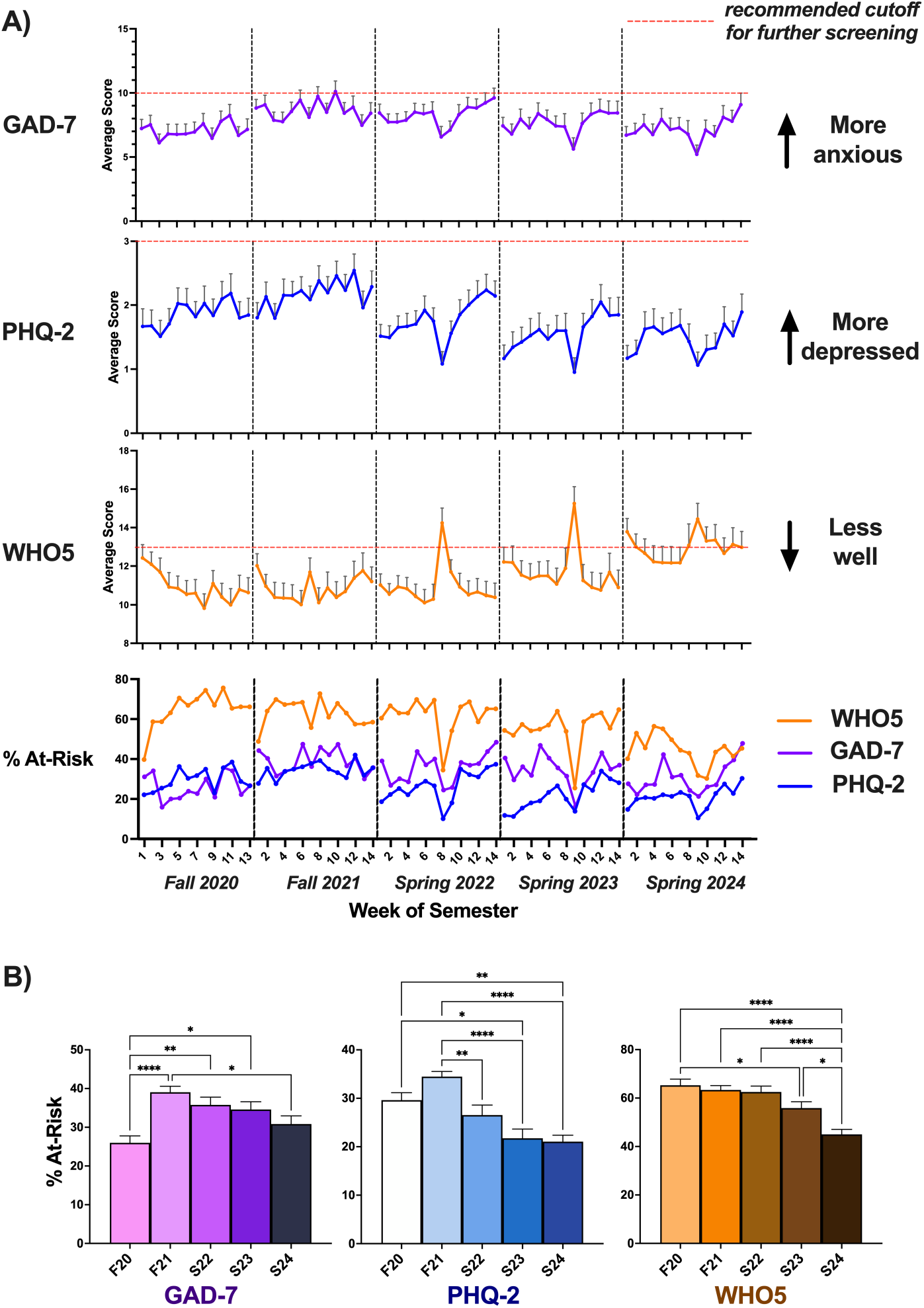
Wellbeing Across Semesters. (**A**) Wellness scale mean scores and percent at-risk over five semesters. *Top three panels:* Average weekly scores are shown for the individual scales, with error bars indicating variability between students at each time point (∼47-69 students were averaged for each data point, depending on the semester; see **Table S1** in the Supplementary Material). *Bottom panel:* For percent at-risk, an aggregate value was obtained at each time point (number of students “at-risk” divided by total students, multiplied by 100). Acute increases in wellbeing were observed around the mid-point of each semester corresponding to the Spring or Fall Break periods. (**B**) Semester averages for percent at-risk, shown for each of the wellness scales. Supporting the trends visible in (A), percent at-risk on the GAD-7 and PHQ-2 peaked in F21. WHO5 percent at-risk values remained relatively stagnant before declining in S23 and falling precipitously in S24. \**P*<..*05*, \*\**P*<..*01*, \*\*\**P*<..*001*, \*\*\*\**P*<..*0001*

#### Overall Descriptive Outcomes

See **Table S1** in the Supplementary Material for the mean percent at-risk each semester for each of the study’s scales. For all three scales, one-way ANOVA revealed a significant effect of semester (GAD-7: *F*_(4, 64)_ = 6.8, *P* =.0001; PHQ-2: *F*_(4, 64)_ = 12.0, *P* <..0001; WHO5: *F*_(4, 64)_ = 13.0, *P* <..0001). To further examine differences between semesters while minimizing Type I error, we applied Tukey’s multiple comparisons tests for each scale (**Figure 1B**).

#### GAD-7

Percent at-risk for anxiety (students scoring ≥ 10 each week) was lowest in F20. The metric significantly increased in F21 (difference, -13.1; 95% CI, -20.69 to -5.43; *P* <..0001), remaining higher than F20 levels in S22 (difference, -9.8; 95% CI, -17.42 to -2.16; *P* =.0054) and S23 (difference, -8.6; 95% CI, -16.22 to -0.97; *P* =.0196) despite gradually decreasing in magnitude. By S24, percent at-risk was reduced from F21 (difference, 8.2; 95% CI, 0.72 to 15.70; *P* = 0.0246) and no different from F20 levels.

#### PHQ-2

Percent at-risk for depression (students scoring ≥ 3 each week) was greatest in F21, then dropped in S22 (difference, 7.9; 95% CI, 1.53 to 14.35; *P* =.0078). Subsequent semesters saw a further reduction, with S23 levels decreased compared to F21 (difference, 12.7; 95% CI, 6.31 to 19.13; *P* <..0001) and F20 (difference, 7.9; 95% CI, 1.34 to 14.41; *P* =.0104), and S24 levels also decreased compared to F21 (difference, 13.4; 95% CI, 7.01 to 19.83; *P* <..0001) and F20 (difference, 8.6; 95% CI, 2.05 to 15.11; *P* =.0042).

#### WHO5

Percent at-risk for low wellbeing (students scoring <.13 each week) did not significantly differ between the first three semesters (F20, F21, and S22). S23 levels were lower than F20 (difference, 9.4; 95% CI, 0.17 to 18.61; *P* =.0438). During S24, percent at-risk was significantly and substantially reduced from F20 (difference, 20.3; 95% CI, 11.05 to 29.49; *P* <..0001), F21 (difference, 18.3; 95% CI, 9.28 to 27.38; *P* <..0001), S22 (difference, 17.5; 95% CI, 8.47 to 26.56; *P* <..0001); and S23 (difference, 10.9; 95% CI, 1.83 to 19.93; *P* =.0107).

### 3.2 Relationship between COVID-19 Levels, Social Restriction Measures, and Wellbeing

We related survey results to local COVID-19 case levels and university social isolation measures. A timeline of major events associated with COVID-19, university Social Gathering Meter changes, and standardized survey scores is shown in **Figure 2**. Correlation analyses were run to assess relationships between exposures and wellness scales (**Figure 3**) or individual survey questions (**Table 1**) at weekly time points.

**Table 1.** Rank-Ordered Correlations between Wellness Scale Questions, COVID-19 Cases, and Social Gathering Restrictions.^a^

| Social Gathering Meter |  | Emory COVID-19 Cases |  | County COVID-19 Cases |  |
| --- | --- | --- | --- | --- | --- |
| Survey Total or Subquestion | Pearson r | Survey Total or Subquestion | Pearson r | Survey Total or Subquestion | Pearson r |
| PHQ-2 Q1 "little interest or pleasure" | **0.470** | GAD-7 Q1 "nervous, anxious" | **0.367** | GAD-7 Q1 "nervous, anxious" | **0.405** |
| PHQ-2 total (Q1-2) | **0.466** | GAD-7 Q2 "can't stop worrying" | **0.276** | GAD-7 Q3 "worrying too much" | **0.288** |
| PHQ-2 Q2 "down, depressed, hopeless" | **0.433** | GAD-7 Q3 "worrying too much" | **0.270** | GAD-7 Q4 "trouble relaxing" | 0.234 |
| GAD-7 Q4 "trouble relaxing" | 0.134 | GAD-7 Q4 "trouble relaxing" | 0.159 | GAD-7 Q2 "can't stop worrying" | 0.233 |
| GAD-7 Q1 "nervous, anxious" | 0.095 | GAD-7 total (Q1-7) | 0.146 | GAD-7 total (Q1-7) | 0.217 |
| GAD-7 Q7 "afraid of something awful" | 0.084 | WHO5 Q5 "filled with interesting things" | 0.145 | GAD-7 Q7 "afraid of something awful" | 0.172 |
| GAD-7 Q6 "annoyed, irritable" | 0.039 | GAD-7 Q7 "afraid of something awful" | 0.111 | PHQ-2 Q2 "down, depressed, hopeless" | 0.130 |
| GAD-7 total (Q1-7) | 0.030 | WHO5 Q1 "cheerful, good spirits" | -0.056 | PHQ-2 total (Q1-2) | 0.063 |
| GAD-7 Q2 "can't stop worrying" | -0.014 | PHQ-2 Q2 "down, depressed, hopeless" | -0.062 | GAD-7 Q5 "restless, hard to sit still" | -0.015 |
| GAD-7 Q5 "restless, hard to sit still" | -0.049 | WHO5 Q3 "active, vigorous" | -0.082 | PHQ-2 Q1 "little interest or pleasure" | -0.021 |
| GAD-7 Q3 "worrying too much" | -0.082 | PHQ-2 total (Q1-2) | -0.125 | GAD-7 Q6 "annoyed, irritable" | -0.069 |
| WHO5 Q2 "calm, relaxed" | **-0.287** | GAD-7 Q5 "restless, hard to sit still" | -0.146 | WHO5 Q5 "filled with interesting things" | -0.106 |
| WHO5 Q1 "cheerful, good spirits" | **-0.383** | WHO5 total (Q1-5) | -0.167 | WHO5 Q1 "cheerful, good spirits" | -0.243 |
| WHO5 Q4 "fresh, rested in AM" | **-0.412** | PHQ-2 Q1 "little interest or pleasure" | -0.182 | WHO5 Q3 "active, vigorous" | **-0.308** |
| WHO5 total (Q1-5) | **-0.536** | GAD-7 Q6 "annoyed, irritable" | -0.251 | WHO5 total (Q1-5) | **-0.360** |
| WHO5 Q3 "active, vigorous" | **-0.603** | WHO5 Q4 "fresh, rested in AM" | **-0.315** | WHO5 Q4 "fresh, rested in AM" | **-0.402** |
| WHO5 Q5 "filled with interesting things" | **-0.611** | WHO5 Q2 "calm, relaxed" | **-0.369** | WHO5 Q2 "calm, relaxed" | **-0.444** |

<sup>a</sup>Significant Pearson r values are shown in bold. For the university Social Gathering Meter, green status was coded as "1" (least restrictive), yellow as "2", modified yellow as "3", and orange as "4" (most socially restrictive). COVID-19 Cases were natural log transformed to deal with outliers.

**Figure 2.**
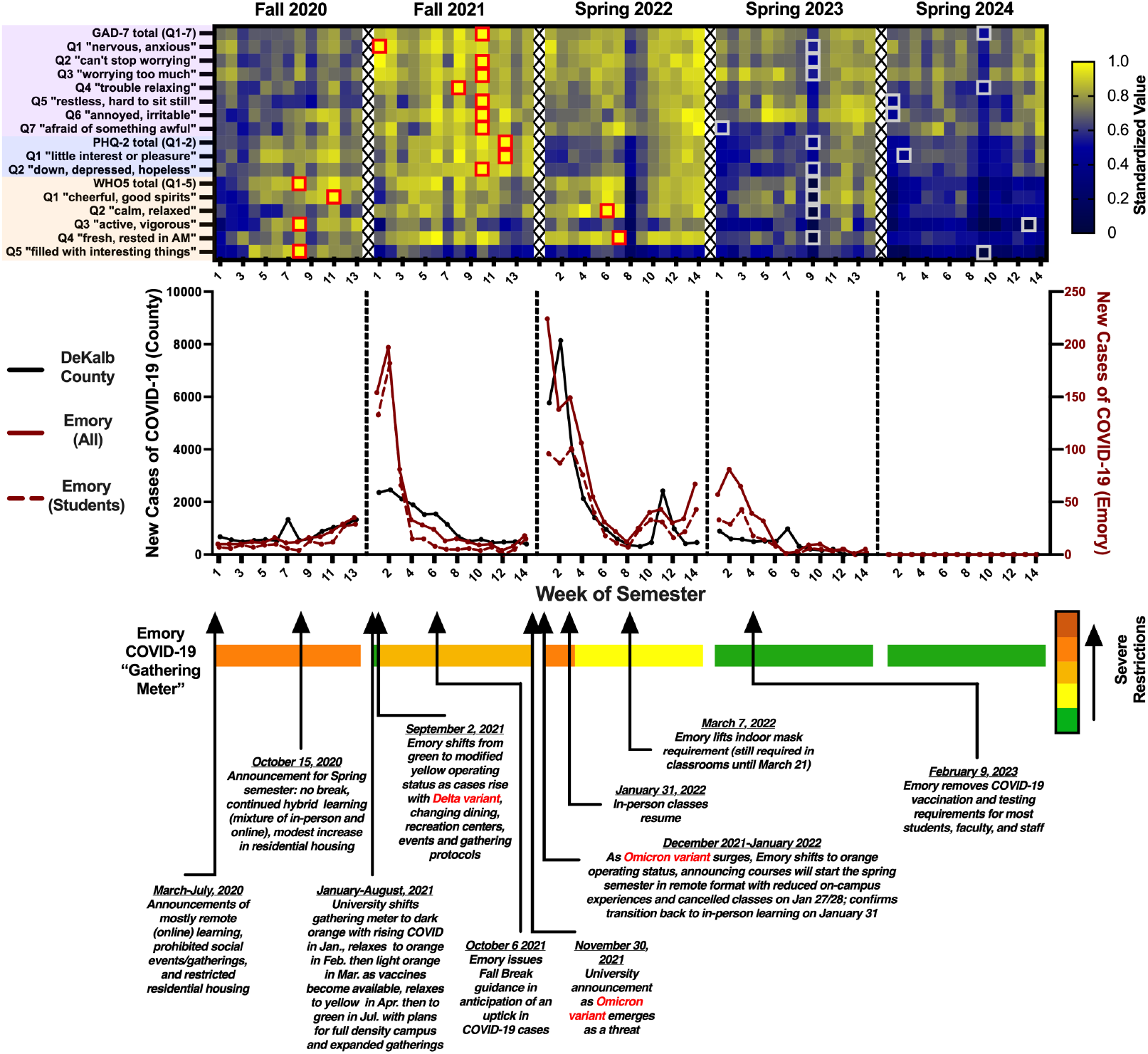
Wellbeing vs. COVID-19 Measures. Wellbeing scales (*top*), COVID-19 case numbers (*middle*), and a timeline of events tied to university social gathering restrictions (*bottom*). Survey responses are shown in heatmap form, with maximal weekly averages for each survey or question standardized to 1.0. WHO5 responses are inverted (higher values indicate worse wellbeing). F20 saw the most severe social gathering restrictions (orange level), during which only small groups of students could gather indoors or outdoors with a faculty or staff sponsor. General wellbeing (WHO5 total) and scores for “active, vigorous”, “filled with interesting things”, and “cheerful, good spirits reached five-year lows during this semester. The gathering meter was updated from green to modified yellow in early F21 due to a rise in COVID cases with the emergence of the Delta variant, corresponding to changes in dining, recreation centers, events and gathering protocols. Nearly all questions from the GAD-7 and PHQ-2 indicated the lowest mental health this semester, as the initial Delta wave subsided and warnings of ongoing isolation persisted with the arrival of Omicron in November. Due to the Omicron variant, S22 classes began in remote format with an uptick to orange status and reduced oncampus experiences, before shifting to yellow with more flexible guidelines for large indoor gatherings. “Calm, relaxed” and “fresh, rested in AM” reached five-year lows coinciding with the tail end of the Omicron wave. In-person learning resumed and the indoor mask requirement was gradually lifted in March. S23 and S24 saw a full removal of restrictions on gathering size, density, and duration. All survey totals and questions indicated peak wellness levels during these semesters. Note that on August 11, 2022, Emory switched to a new, two-level campus operating model (standard vs. heightened response); we depict “standard” as the green condition for simplicity.

**Figure 3.**
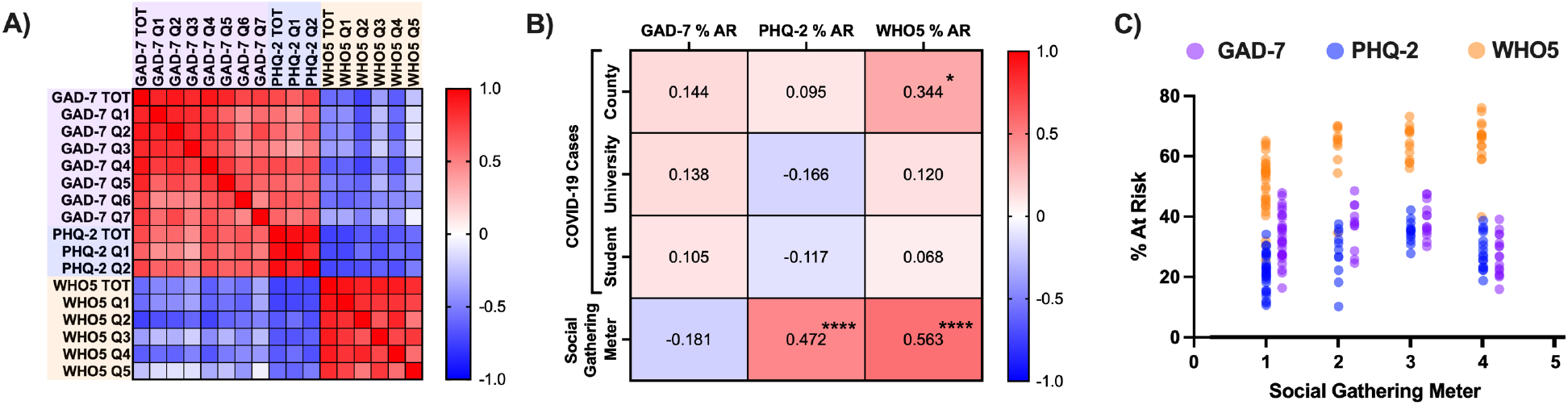
Correlations between COVID-19, Social Restrictions, and Wellbeing. Associations between weekly survey results, COVID-19 case levels, and the university social gathering meter were investigated using Pearson correlation analysis. COVID-19 data were natural log-transformed before analysis to exclude outliers (e.g. COVID-19 case levels on early weeks of semesters). (**A**) Intra- and inter-scale reliability. There were generally strong associations between student responses on individual questions both within and across surveys, and between survey totals (see Results). (**B**) Weekly COVID-19 case levels and social restrictions versus wellness scales. Increased COVID-19 levels at the county level predicted greater % at-risk (AR) for poor wellbeing and more severe social restrictions predicted greater % AR for depression and poor wellbeing. (**C**) Increasingly restrictive social gathering measures (higher numbers = more restrictions) strongly predicted risk for depression and poor wellbeing, but not anxiety. Individual dots represent weekly averages for a particular combination of survey scores and social gathering metrics. The individual basis of highlight correlations shown in (B) is clearly visible. \**P*<..*05*, \*\**P*<..*01*, \*\*\**P*<..*001*, \*\*\*\**P*<..*0001*

#### Inter- and intra-scale reliability (Figure 3A)

Pearson r values were tabulated to determine whether total scores were correlated between scales, and relationships between individual questions within a given scale. As expected, weekly GAD-7 averages were significantly positively correlated with PHQ-2 averages (r = 0.715, *P* <..0001) and negatively correlated with WHO5 averages (r = -0.581, *P* <..0001), and weekly PHQ-2 averages were negatively correlated with WHO5 averages (r = -0.746, *P* <..0001). Within the GAD-7, correlations between individual questions ranged from to 0.455 (Q1 “nervous, anxious” and Q6 “annoyed, irritable”) to 0.863 (Q1 “nervous, anxious” and Q2 “can’t stop worrying”). Within the PHQ-2, the correlation between questions Q1 (“little interest or pleasure”) and Q2 (“down, depressed, hopeless”) was 0.828. Within the WHO5, correlations ranged from 0.533 (Q2 “calm, relaxed” and Q5 “filled with interesting things”) to 0.860 (Q2 “calm, relaxed” and Q4 “fresh, rested in AM”).

#### Correlations between survey percent at-risk and exposures

Pearson r values were tabulated to determine the relationship between weekly survey results (percent at-risk), social gathering measures, and local COVID-19 case numbers. Values are shown in **Figure 3B**, with highlighted correlations displayed graphically in **Figure 3C**. Social gathering restrictions were strongly positively correlated with percent at-risk for depression (r = 0.47, *P* = 4.3E-05) and poor wellbeing (r = 0.56, *P* = 4.8E-07) but showed a trend toward negative correlation with anxiety (r = -0.18, *P* = 0.14), which was significant when Spring Break weeks were excluded (r = -0.26, *P* = 0.036). In contrast, COVID-19 case numbers at the county level were positively correlated with percent at-risk for poor wellbeing (r = 0.34, *P* = 0.013). No other correlations reached significance.

#### Correlations between survey means and exposures

Pearson r values were tabulated to determine the relationship between overall and individual survey question responses (weekly means), social gathering measures, and local COVID-19 case numbers (**Table 1**). The strongest positive correlate of more severe social gathering meter restrictions was “little interest or pleasure” (PHQ-2 Q1) (r = 0.47, *P* = 4.5E-05) while the strongest negative correlate was “filled with interesting things” (WHO5 Q5) (r = -0.61, *P* = 2.5E-08). The strongest positive correlate of local COVID-19 cases was “nervous, anxious” (GAD-7 Q1) (Emory students, faculty, and staff: r = 0.37, *P* = 0.0059; Dekalb County: r = 0.40, *P* = 0.0029) while the strongest negative correlate was “calm, relaxed” (WHO5 Q2) (Emory students, faculty, and staff: r = -0.37, *P* = 0.0055; Dekalb County: r = -0.44, *P* = 0.00099).

## 4 Discussion

### 4.1. Mental Health Across Semesters

The COVID-19 pandemic coincided with a mental health crisis that plagued college campuses nationwide (13-15). We collected data on student wellbeing, local COVID-19 prevalence, and social isolation measures at weekly time points from 277 university students over five semesters from 2020-2024. Survey responses indicated increased depression and anxiety and reduced wellbeing in the midst of the COVID-19 pandemic (2020-2022), with an improvement in these metrics as concerns about infection waned and social restrictions were terminated during the 2023-2024 semesters. The most severe social isolation measures, implemented in F20, preceded a precipitous decline in student mental health (**Figures 1 and 2**). Anxiety (measured via the GAD-7) and depression (measured via the PHQ-2) spiked in F21 as the Delta variant of COVID-19 caused prolonged disruptions. Amidst the ongoing uncertainty sandwiched between the Delta and Omicron variants (with Omicron emerging in late November 2021), responses to nearly all of the GAD-7 and PHQ-2 questions reached their peaks. General wellbeing (measured via the WHO5), in contrast, declined quickly the first few weeks of F20 and languished at a trough from F20 through S22, with a high percentage of students considered at-risk for poor wellbeing during all three semesters. At the tail-end of the Omicron wave in mid-S22, immediately preceding the university’s announcement of reinstated in-person learning and removal of mask restrictions, students rated themselves the least fresh, rested, and relaxed of any point over the five years, possibly reflecting sleep difficulties (3) or a state of burnout (16) associated with the toll of an extended period of chronic unpredictable stress. Despite a return to a more normalized social environment, wellbeing scores lagged behind, trending upward only in S23 and peaking in S24.

### 4.2. Associations Between COVID-19 Prevalence, Social Isolation, and Mental Health

To better understand pandemic-related factors that might predict specific syndromal patterns, we evaluated the separate associations of social restrictions and infection prevalence with PHQ-2, GAD-7 and WHO5 scores at weekly time points. Positive associations were observed between severity of university social gathering restrictions and risk for both depression and reduced wellbeing, but not anxiety (**Figure 3**). Analyses of individual items confirmed that every item from the PHQ-2 and WHO5 scales was significantly correlated with social gathering meter levels, whereas none of the anxiety items were (**Table 1**). In contrast, item analysis revealed that increased infection prevalence was associated with heightened anxiety and reduced wellbeing, but not with increases in depression. There were especially robust associations between COVID-19 infection rates and anxiety, and social restriction measures and increases in items related to anhedonia. Since the highest COVID-19 values were almost always observed near the start of a semester (**Figure 2**), when students typically report better mental health, the strength of our observed correlations may have been reduced. Nonetheless, these findings provide information that will be important for considering how to best manage the implications of future pandemics for college student mental health. While social restrictions may always be important for disease containment, identifying and implementing strategies for reducing the hedonic costs of these measures will be essential.

### 4.3. Study Limitations

The current study has several important limitations. Students self-selected to enroll in our university course. Students already facing or more prone to develop mental health challenges may be more likely to enroll in a course on mental wellbeing and resilience. Furthermore, the GAD-7, PHQ-2, and WHO-5 were designed for measuring generalized anxiety disorder (17), depression (18, 19), and wellbeing (20), respectively, in the general population rather than specifically targeting young adults or university students. Nonetheless, percent at-risk levels were similar to those observed in other student cohorts.(6) Because two of the cohorts filled out the surveys in the fall (F20/F21) and three in the spring (S22-24), changes in wellbeing due to seasonal effects or the structure of an academic year (which typically runs from late August to the following May) may pose a potential confound for the interpretation of our findings. For example, students taking the class in the spring may anticipate the upcoming summer break with enthusiasm. Alternatively, seasonal depression often occurs in the late fall or early winter, possibly leading to lower fall semester wellness scores. While this confound cannot be entirely ruled out, S22 was one of the worst two semesters for student mental health despite an emerging return to normalcy. If seasonal effects were important, one might also predict differential trajectories in wellbeing over the course of a fall and spring semester, i.e. gradual deterioration of mental health in the fall and a rise in the spring. However, this was not the case (see **Figure 1**), as across all cohorts mental health declined from the start to the end of each semester.

### 4.4. Conclusions

The COVID-19 pandemic took a heavy toll on the mental wellbeing of our university undergraduates, with signs of anxiety as local COVID-19 prevalence skyrocketed and signs of anhedonia as social gatherings were restricted. While mental health gradually recovered with the return to normalcy, at the final time point data was collected (Week 14 of S24), around 30-40% of students remained at-risk for depression or anxiety based on survey guidelines. Even with the pandemic waning, Americans increasingly spend time alone (21). Clearly, student wellbeing remains a pressing cause for concern and there is ample room for novel interventional and educational research.

## Author Contributions

DJN and CLR formulated the research question and designed and carried out the study. DJN analysed the data and wrote the article, with substantial feedback and revisions from CLR.

## Data Availability Statement

These student datasets cannot be shared for legal, ethical, or privacy reasons.

