## Supplementary Material for "Tracking Student Wellbeing Throughout the COVID-19 Pandemic"

### 1 Table S1. Class Compositions, University COVID-19 Cases, and Percent At-Risk Values

| | F20 | F21 | S22 | S23 | S24 | AVG/TOT[a] (% of full data set) |
|---|---|---|---|---|---|---|
| **Class Composition** | | | | | | |
| Freshman (% of class) | 3 (6.4) | 4 (6.2) | 13 (18.8) | 1 (2.1) | 0 (0) | 21 (7.6) |
| Sophomore (% of class) | 17 (36.2) | 19 (29.2) | 11 (15.9) | 14 (29.8) | 12 (24.5) | 73 (26.4) |
| Junior (% of class) | 15 (31.9) | 22 (33.8) | 10 (14.5) | 10 (21.3) | 20 (40.8) | 77 (27.8) |
| Senior (% of class) | 12 (25.5) | 20 (30.8) | 35[b] (50.7) | 22 (46.8) | 17 (34.7) | 106 (38.3) |
| Total Enrolled (% of full data set) | 47 (17.0) | 65 (23.5) | 69[b] (24.9) | 47 (17.0) | 49 (17.7) | 277 |
| **Local COVID-19 Cases** | | | | | | |
| Dekalb County (% of full data set) | 10,179 (16.8) | 16,603 (27.4) | 28,368 (46.8) | 5,426[d] (9.0) | No data | 60,576 |
| Emory Total[c] (% of full data set) | 211 (10.0) | 606 (28.6) | 976 (46.1) | 323 (15.3) | No data | 2,116 |
| Emory Students (% of full data set) | 149 (10.7) | 466 (33.6) | 605 (43.6) | 168 (12.1) | No data | 1,388 |
| **% At-risk for Poor Mental Health** | | | | | | |
| GAD-7 (mean ± SD) | 25.99 ± 6.40 | 39.04 ± 5.78 | 35.78 ± 7.44 | 34.58 ± 7.52 | 30.83 ± 7.87 | 33.24 |
| PHQ-2 (mean ± SD) | 29.62 ± 5.52 | 34.47 ± 4.02 | 26.52 ± 7.72 | 21.75 ± 7.11 | 21.05 ± 5.02 | 26.68 |
| WHO5 (mean ± SD) | 65.28 ± 9.16 | 63.34 ± 6.72 | 62.52 ± 9.11 | 55.89 ± 9.66 | 45.01 ± 7.70 | 58.41 |

[a]***AVG/TOT = average and total***
[b]***2 Seniors included in total withdrew***
[c]***Includes students, faculty, and staff***
[d]***County case numbers were not reported the last three weeks of S23***

### Table S2. Survey Instrument[a]

**GAD-7:** "Over the **last week**, how often have you been bothered by the following problems?"

*0 — Not at all; 1 — Several days;*
*2 — More than half the days; 3 — Nearly every day*

**Q1. Feeling nervous, anxious, or on edge**
**Q2. Not being able to stop or control worrying**
**Q3. Worrying too much about different things**
**Q4. Trouble relaxing**
**Q5. Being so restless that it's hard to sit still**
**Q6. Becoming easily annoyed or irritable**
**Q7. Feeling afraid as if something awful might happen**

*Recommended Cutoff:* **10 or greater**

**PHQ-2**: "Over the **last week**, how often have you been bothered by the following problems?"

*0 — Not at all; 1 — Several days;*
*2 — More than half the days; 3 — Nearly every day*

**Q1. Little interest or pleasure in doing things**
**Q2. Feeling down, depressed, or hopeless**

*Recommended Cutoff:* **3 or greater**

**WHO5:** "Please indicate how you have been feeling over the **last week** for each of the five statements."

*0 — At no time; 1 — Some of the time; 2 — Less than half of the time;*
*3 — More than half of the time; 4 — Most of the time; 5 — All of the time*

**Q1. I have felt cheerful and in good spirits**
**Q2. I have felt calm and relaxed**
**Q3. I have felt active and vigorous**
**Q4. I woke up feeling fresh and rested**
**Q5. My daily life has been filled with things that interest me**

*Recommended Cutoff:* **below 13**

[a]***Scales were administered in the Canvas Learning Management System; students used dropdown menus to select scores for each question.***

### 3 Results S1. Survey Means

Overall Descriptive Outcomes: Across cohorts, the mean (SD) total scores for each scale were 7.8 ± 1.0 (GAD-7, out of 21), 1.8 ± 0.3 (PHQ-2, out of 6) and 11.5 ± 1.2 (WHO5, out of 25). Anxiety, depression, and wellbeing all indicated poorest mental health during the F21 semester, with average scores of 8.7 ± 0.8 (GAD-7), 2.2 ± 0.2 (PHQ-2) and 10.9 ± 0.6 (WHO5). By S24, scores improved to 7.2 ± 0.9 (GAD-7), 1.5 ± 0.2 (PHQ-2) and 12.9 ± 0.7 (WHO5). For all three scales, one-way ANOVA revealed a significant effect of semester (GAD-7: $F_{(4, 64)} = 10.3$, $P < .0001$; PHQ-2: $F_{(4, 64)} = 15.8$, $P < .0001$; WHO5: $F_{(4, 64)} = 14.7$, $P < .0001$). To further examine differences between semesters while minimizing Type I error, we applied Tukey's multiple comparisons tests for each scale.

GAD-7: Anxiety levels rose significantly from F20 to F21 (difference, -1.6; 95% CI, -2.41 to -0.72; $P < .0001$) and S22 (difference, -1.2; 95% CI, -2.02 to -0.33; $P = .0020$), whereas they did not differ between F20 and the later semesters (S23/24). Anxiety scores decreased from F21 to S23 (difference, 0.97; 95% CI, 0.14 to 1.79; $P = .0137$) and further decreased in S24 to drop below F21 (difference, 1.4; 95% CI, 0.62 to 2.27; $P < .0001$) and S22 (difference, 1.1; 95% CI, 0.23 to 1.88; $P = .0058$) levels.

PHQ-2: Depression levels rose significantly from F20 to F21 (difference, -0.31; 95% CI, -0.58 to -0.036; $P = .0188$) and then dropped from F21 to S22 (difference, 0.41; 95% CI, 0.14 to 0.68; $P = .0007$) such that there was no difference between F20 and S22. Depression scores fell further in S23 from F20 (difference, 0.30; 95% CI, 0.021 to 0.57; $P = .0283$) and F21 (difference, 0.61; 95% CI, 0.34 to 0.88; $P < .0001$) and were significantly decreased in S24 from F20 (difference, 0.37; 95% CI, 0.10 to 0.65; $P = .0026$), F21 (difference, 0.68; 95% CI, 0.42 to 0.95; $P < .0001$), and S22 (difference, 0.28; 95% CI, 0.0093 to 0.55; $P = .0391$) levels.

WHO5: There were no differences in average WHO5-assessed wellbeing between the first four semesters. In contrast, scores increased significantly in S24 compared to all prior semesters ($P < .0001$ vs. F20, F21, and S22; $P = .0033$ vs. S23).
